# Development and Preliminary Evaluation of the Patient–Artificial Intelligence Rating Instrument

**DOI:** 10.64898/2026.10.20.26366053

**Authors:** Adam Rodman, Elizabeth Lowe, Eric J. Thomas, Yoel Benarroch, Long Ngo, Talya Salant, Fabienne C. Bourgeois, Mary Newton, Chase Walton, Sigall K. Bell

**Affiliations:** Patient and Family Advisory Council, Beth Israel Deaconess Medical Center, Boston, Massachusetts; Department of Internal Medicine, McGovern Medical School, The University of Texas Health Science Center at Houston (UTHealth Houston), Houston, Texas; UTHealth Houston–Memorial Hermann Center for Healthcare Quality and Safety, Houston, Texas; Department of Medicine, Beth Israel Deaconess Medical Center, Boston, Massachusetts; Harvard Medical School, Boston, Massachusetts; Boston Children’s Hospital, Boston, Massachusetts; Center of Surgical and Medical Intelligence, Beth Israel Lahey Health, Boston, Massachusetts; Bowdoin Street Health Center, Dorchester, Massachusetts; Division of Hospital Medicine, Beth Israel Deaconess Medical Center, Boston, Massachusetts; Beth Israel Deaconess Medical Center, Boston, Massachusetts

## Abstract

**Importance:** Patients increasingly use artificial intelligence (AI) for health-related conversations, but evaluation measures largely reflect clinician judgment.

**Objective:** To develop and preliminarily evaluate a patient instrument assessing the quality of patient–AI health conversations.

**Design:** Cross-sectional survey conducted June 5-14, 2026, following patient-partnered instrument development and cognitive interviews.

**Setting:** An anonymous online survey.

**Participants:** A convenience sample of adults with prior use of large language models for personal health-related queries, recruited from an academic medical center-affiliated Insight Panel and patient peers.

**Exposures:** Participants’ own prior health-related conversations with AI tools of their choosing.

**Main Outcomes and Measures:** The Patient-AI Rating instrument comprised 12 communication-quality and patient engagement items rated from 1 to 5 and a separately scored patient-perceived-harm item. Internal consistency was assessed using Cronbach α, with sensitivity analyses using first ratings only for those using the tool to rate multiple conversations and participant-mean profiles. We examined item–total correlations, inter-item structure, and between- and within-participant variation.

**Results:** Fourteen participants contributed 46 complete ratings; among 13 providing demographics, median age was 63 years (IQR, 53-70 years), and 8 (62%) reported female gender. Participants provided a median of 2 ratings (range, 1-17). Cronbach α was 0.88 (95% CI, 0.83-0.93) across all ratings, 0.85 using first ratings only, and 0.92 using participant-mean profiles. Corrected item–total correlations ranged from 0.42 to 0.72; mean inter-item correlation was 0.40. No item deletion improved Cronbach α. The mean quality score was 4.43 (SD, 0.45). Among 9 participants contributing multiple ratings, the intraclass correlation coefficient was 0.45, suggesting inter-conversation variability. No perceived harm was reported in 44 of 46 ratings (96%); the harm item correlated minimally with the instrument total (r = 0.09).

**Conclusions and Relevance:** In this small cross-sectional study of engaged patients, the Patient-AI Rating instrument showed promising internal consistency and variation across rated conversations. These findings support further evaluation of patient-reported communication quality alongside clinician-based measures. Validation in larger, more diverse populations, including assessment of construct and criterion validity, is needed before broader use.

**Key Points:** *Question:* Can a patient instrument measure the quality of patients’ health-related conversations with artificial intelligence (AI)?

*Findings:* In this cross-sectional study, 14 participants provided 46 ratings of their health-related AI conversations using a 12-item scale and a perceived-harm item (the Patient AI Rating (PAIR) instrument). The patient metric showed good internal consistency, and scores varied both by raters and across conversations rated by the same participant.

*Meaning:* The PAIR instrument offers a preliminary approach to incorporate patient perspective and experience into AI evaluation – a measure largely missing from current AI assessment; larger studies are needed to establish validity and generalizability.

## Introduction

As large language models rapidly proliferate in healthcare, measurement science of LLM-generated text has rapidly matured. The outputs from AI scribes are routinely audited with validated psychometric instruments such as the Provider Documentation Summarization Quality Instrument^1^ or rubric-based frameworks.^2^ Multiple blinded studies have now compared LLM- and physician-generated discharge summaries,^3^ and increasingly researchers are using emerging error taxonomies that classify hallucination, omission, and potential for patient harm.^4^ However, virtually every one of these advances in measurement science share a fundamental assumption – physicians and their values are the arbiter of the quality of healthcare-related LLM outputs.

While at least partially justifiable when tools primarily face physicians, patient-facing LLMs are now a significant part of the health ecosystem. Approximately one-third of American adults now routinely use LLMs to better understand their own health,^5^ and this activity largely takes place outside of healthcare contexts. While patient-facing systems have demonstrated impressive clinical abilities, physician-judged quality and patient experience are distinct constructs.^6^ Surveys of clinicians and patients have found that clinicians are substantially more receptive than patients to AI in patient-facing roles, and that providers rank patient communication as the least important role of clinical documentation.^7^ Patients consistently name priorities — transparency, empathy, personalization, language concordance — that are not routinely captured in physician-anchored AI metrics.^7,8^ An AI text output can be accurate, complete, and physician-approved, and still fail to be understood, trusted, or usable by a patient. Measurement science requires a new paradigm: integrating patient-experienced quality of AI health communication. As an example, a recent study investigated the utility of ChatGPT Health for patient-facing triage of potentially life-threatening concerns.^9^ The authors used sixty clinician-authored vignettes and used a ground truth of physician consensus on the appropriateness of AI judgment as the outcome. No patient-derived measures were used to study the outcomes of a tool that directly faces (and affects) patients – not because of an error from the investigators, but because such a measure does not exist. Yet patients have these real unmeasured conversations about triage decisions in their homes every day.

As patient-facing AI transitions from ad hoc individual use toward health-system level deployment, there is a need for patient-derived measures of quality. Here we report the development, with patient and community partners, of the preliminary psychometric evaluation of the Patient-AI Rating (PAIR) instrument. To our knowledge, PAIR is the first patient-reported measure of the quality of AI-generated health communication.

## Methods

### Instrument development

The PAIR instrument was developed through a multi-stage, patient-partnered process. We began with a review of the published literature on the evaluation of generative AI in health communication and on patient perspectives toward AI in healthcare. This review encompassed three bodies of work. First, validated instruments for rating AI-generated clinical text — including the Provider Documentation Summarization Quality Instrument (PDSQI-9)^1^ and rubric-based evaluations of ambient digital scribe platforms^2^ — which assess dimensions such as accuracy, completeness, usefulness, comprehensibility, and stigmatizing language, though exclusively from the clinician perspective. Second, we reviewed studies of the quality and safety of AI-generated clinical text, including blinded comparison of LLM- and physician-generated discharge summaries,^3^ a preliminary patient-safety error classification for generative AI,^4^ and harm-scale–based analyses identifying omissions, inaccuracies, hallucinations, and potential for patient harm as recurring failure modes of LLM output.^2–4^ Third, we reviewed studies of patient and caregiver perspectives on AI in healthcare, including small-group and survey studies of generative AI in patient-centered clinical decision support^8^ and in patient-accessible clinical documentation,^7^ together with primary studies and systematic and scoping reviews of patient preferences for AI in healthcare spanning both general populations^10–16^ and patients with limited English proficiency.^17–20^ Across this third body of work, we identified recurring patient-identified priorities included transparency and disclosure of AI use,^10,11,14^ accuracy and human oversight,^7,11,19^ preservation of the human relationship and empathic communication,^8,11^ plain language and accessibility across literacy levels and languages,^17,18,20^ freedom from bias and culturally insensitive output,^12,17,19^ personalization,^8^ and patient choice and engagement in AI development.^10,13^ Our review confirmed that while multiple validated measures existed for evaluating AI output from the physician perspective, no validated patient-reported measure of AI health-communication quality was available.

Working with a patient-researcher, the study team extracted from the reviewed literature a list of candidate domains potentially relevant to patients’ assessment of AI-generated health conversations, drawing both on the quality dimensions of clinician-facing instruments (e.g., accuracy, completeness, usefulness, comprehensibility, harm, stigmatizing language)^1–4^ and on the patient-identified priorities above (e.g., empathy, personalization, language and reading level, respectfulness).^7,8,10–20^ This candidate list was then iterated with members of two patient and community advisory groups: the BIDMC Patient and Family Advisory Council (PFAC) and the Community Advisory Board of Bowdoin Street Health Center, a BIDMC-affiliated community health center. We asked patients what they thought would be important to measure in terms of AI’s performance and how they would know (or measure) whether a conversation with AI was good. After discussion with members of these groups, we constructed a draft rating instrument and definitions for each item together with our patient-researcher.

The draft instrument was then refined through cognitive interviews with four patients including members of the PFAC, Community Advisory Board, and the leader of a national patient safety patient advocacy group who interacts regularly with hundreds of patients as part of her work on patient-reported measure prioritization. Each gave direct feedback on the instrument. The interviews included four steps: 1) item-by-item review for understandability, meaning, and whether responses elicited the intended information (“repeat the item in your own words,” “what do you think of when your read this item,” “how would you answer this question”; 2) Item prioritization, including any recommendations for removal of candidate items judged redundant or of low relevance (one item: empowering) and elicitation of any new domains that were perceived by patients as important but missing (none); 3) Review and simplification of item language and domain definitions so that each item captured a single, distinct construct in plain language; and 4) Overall instrument ease of use, flow/order, terminology, and length. This iterative process yielded the final 13-item instrument: twelve items assessing dimensions of communication quality and one item assessing perceived harm.

### Instrument

The Patient-AI Rating (PAIR) instrument is a 13-item patient-reported measure of the quality of AI-generated health communication. Twelve items assess distinct dimensions of information quality and patient engagement: understandability, usefulness, preferred language and reading level, respectfulness, empathy/tone, completeness, accuracy, personalization, focus on health issues that matter most to the patient, contribution to understanding one’s health, preparation for future care, and support for informed decision-making. Each item is rated on a 5-point Likert scale (1 = “Not at all” to 5 = “Extremely”). Item 8 assesses perceived harm on the same response scale and was designed a priori as a distinct safety-screening construct rather than an indicator of communication quality; it is scored and reported separately from the other 12-items on the quality scale. Four items (assessing disrespect or bias, missing information, possible errors, and harm) include optional free-text follow-up prompts to elicit qualitative detail. Participants responded to the instrument shown in Figure 1. The conversations they rated were with AI tools of their choice.

**Figure 1.**
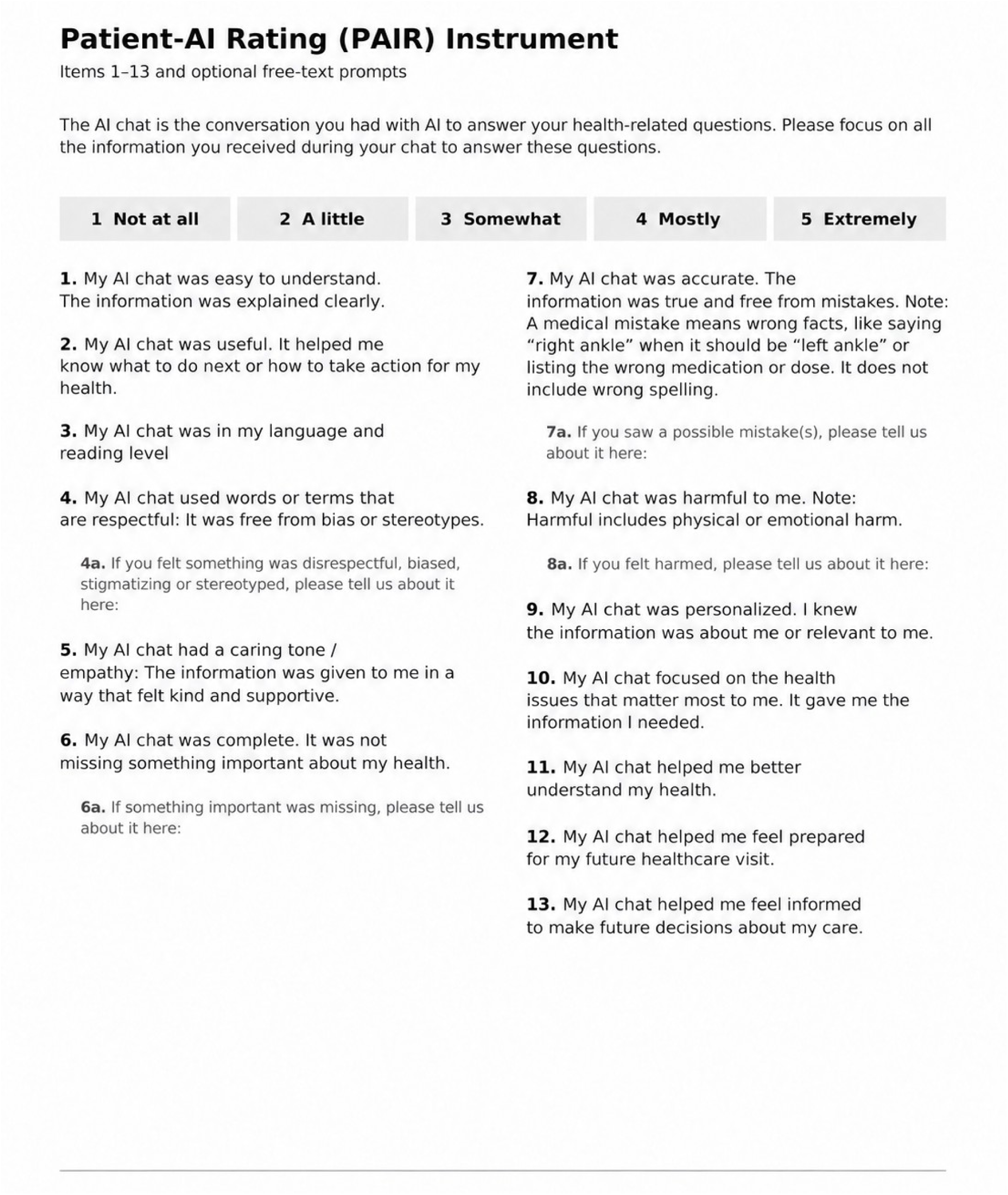
Patient-AI Rating Instrument.

### Participants and setting

We conducted a cross-sectional survey study in which participants rated one or more of their own prior AI health-related conversations. Participants were members of an Insight Panel, an informal community of individuals affiliated with BIDMC, or other patient peers, who are active users of generative AI tools for their own health information needs. Eligible participants were adults (age ≥18 years) with prior use of a large language model for personal health-related queries. Members were invited by an initial recruitment email — distributed through the OpenNotes Insight Panel email list and through peer patient contacts of the study’s patient-researcher — describing the study, its voluntary nature, and the process; prospective agreement was established by reply before any study activity. Consenting participants then received an anonymous Qualtrics survey link, which they could complete in a setting of their choosing. Per protocol, participants could rate multiple different AI-generated health conversations from their own prior use, typically accessed through the saved chat history of their AI tool.

Participants completed the PAIR instrument once for each unique conversation; each rating was submitted through the anonymous survey link as a separate response. Unique participant identifiers were used to link repeated ratings from the same participant. No protected health information or directly identifying data were collected in the survey. Participants were compensated for their time. Ratings were collected between June 5 and June 14, 2026. The BIDMC Committee on Clinical Investigations determined the rating study to be exempt (2025P000873). Instrument development, including cognitive interviews, was conducted under a separate approved protocol (2026P000116). Participants indicated agreement by email before receiving the survey link. This study followed the Strengthening the Reporting of Observational Studies in Epidemiology (STROBE) reporting guideline.

### Statistical analysis

No formal sample-size calculation was performed; this was a preliminary evaluation and we invited all eligible panel members, with sample size determined by panel size and response during the collection window. Item responses were scored 1–5 from labeled response options (not at all to extremely). Because participants could rate multiple AI conversations, we analyzed the data at the level of individual chats, accounting for repeat users (clustering) in our analysis. Internal consistency of the 12-item scale was estimated with Cronbach’s coefficient alpha computed across all chat ratings, with 95% confidence intervals computed by the method of Feldt. To address the potential impacts of clustering, we recomputed the Cronbach’s alpha using (a) only each participant’s first chronological rating and (b) participant-mean item profiles. To characterize the contribution of between- and within-participant variation, we partitioned the variance of the 12-item scale score with a one-way random-effects analysis of variance across participants contributing two or more ratings, from which we report the intraclass correlation coefficient, ICC(1), and the within-participant standard deviation of scale scores across chats. We examined item-level performance using item means and standard deviations, corrected item–total correlations (each item correlated with the sum of the remaining scale items), and alpha-if-item-deleted, computed across all ratings. Inter-item structure was characterized with the full inter-item covariance and correlation matrices, including the mean inter-item correlation. The harm item was summarized descriptively as a frequency distribution at both the rating and participant level. Only complete PAIR ratings were included in psychometric analyses. Demographic and AI-use data used available responses; skipped responses were not imputed. The investigators selected the statistical analyses, and Claude Fable 5 (Anthropic) assisted in developing the code. Analyses were executed in Python (version 3.10) using pandas, NumPy, and SciPy, with human review of the AI-assisted output. All computational analyses were reviewed with a biostatistician.

## Results

### Sample

Fourteen participants contributed 46 completed PAIR ratings of AI-generated health conversations (median, 2 ratings per participant; range, 1–17). Demographic data were optional and were available for 13 of the 14 participants (Table 1). Five participants rated a single conversation, and nine rated two or more. All 46 ratings contained complete responses to all 13 items and were included in the analysis. Four additional incomplete survey submissions were excluded.

**Table 1.** Participant Characteristics.

| **Characteristic** | **Participants, No. (%)**<br>**(n = 13)** |
| --- | --- |
| **Age, years, median (IQR)** | 63 (53–70) |
| **Gender** |  |
| Female | 8 (62) |
| Male | 4 (31) |
| Non-binary | 1 (8) |
| **Race** |  |
| White | 7 (54) |
| Black or African American | 4 (31) |
| Native American | 1 (8) |
| Asian | 1 (8) |
| **Ethnicity** |  |
| Hispanic or Latino | 0 (0) |
| Non-Hispanic or Latino | 13 (100) |
| **Highest level of education** |  |
| Less than or including high school completion | 1 (8) |
| Some college | 3 (23) |
| College graduate (4-year) | 4 (31) |
| Graduate or professional degree | 5 (38) |
| **Primary language** |  |
| English | 12 (92) |
| Spanish | 1 (8) |
| **Confidence completing medical forms in English** |  |
| Extremely | 11 (85) |
| Quite a bit | 1 (8) |
| Somewhat | 1 (8) |
| **Digital literacy** (agree or strongly agree) |  |
| Can use applications (e.g., Zoom) independently | 10 (77) |
| Can set up a video chat independently | 10 (77) |
| Can solve basic technical issues independently | 10 (77) |
| **Has used AI for own health** | 11 (100) |
| **Patient portal use** |  |
| Uses patient portal<sup>a</sup> | 9 (69) |
| Used portal in past 12 months | 8 (62) |
| Used portal in past 6 months | 7 (54) |
| **Has a family member, friend, or care partner** who helps with medical appointments or health information | 3 (23) |

Abbreviations: AI, artificial intelligence; IQR, interquartile range. Values are No. (%) unless otherwise indicated. Demographics were available for 13 of 14 participants. For AI use for one's own health, the denominator is 11: 2 of the 13 participants skipped this question. Percentages may not sum to 100 because of rounding. <sup>a</sup> Patient portal use indicates whether the participant had logged into Epic MyChart.

### Use types

Patient-AI health chats focused on understanding a laboratory (n=14, 30%), radiology or procedure (n=16, 35%) result; preparing for a health visit (n=15, 33%); and/or a diagnostic inquiry (n=17, 37%, Table 2). Twenty (44%) of chats aimed to identify questions to ask clinicians and 28 (61%) were general health inquiries. For these chats, patients entered their notes (n=16, 35%), laboratory (n=15, 33%) or radiology (n=13, 28%) results and/or symptoms (n=15, 33%) into the AI for these chats (Table 2).

**Table 2.** Purposes of Conversations and Health Information Entered Into Artificial Intelligence Tools.

| Characteristic | No. | % |
| --- | --- | --- |
| **Main purpose of the conversation** |  |  |
| Answer a specific question about my health condition | 28 | 61 |
| Questions I should ask my provider | 20 | 44 |
| See what could be causing my symptoms or test results | 17 | 37 |
| Understand my radiology or procedure result | 16 | 35 |
| Prepare for my next visit | 15 | 33 |
| Understand my lab result | 14 | 30 |
| Learn more about my health in general | 6 | 13 |
| Remind me what I was supposed to do after my last visit | 5 | 11 |
| Understand or review my clinician's notes | 4 | 9 |
| Review medications, side effects, or allergies | 4 | 9 |
| Help care for a family member | 3 | 7 |
| Other (decide whether to go to the emergency department)<sup>b</sup> | 1 | 2 |
| **Health information entered into the AI tool** |  |  |
| My notes (or part of my notes) | 16 | 35 |
| My lab result(s) | 15 | 33 |
| My symptoms (no direct information from my health record) | 15 | 33 |
| My radiology or procedural report (or part of my report) | 13 | 28 |
| Medications (own or family member's) | 7 | 15 |
| Self-reported medical history or diagnosis | 2 | 4 |
| Did not enter any personal health information from my record | 5 | 11 |

Abbreviation: AI, artificial intelligence. From 46 rated conversations from 14 participants; some participants contributed more than one rating.

### Internal consistency

Across all 46 chat ratings, the 12-item scale demonstrated good internal consistency, with Cronbach’s alpha of 0.88 (95% CI, 0.83–0.93). The Cronbach’s alpha was 0.85 (95% CI, 0.71–0.94) when restricted to each participant’s first rating (n = 14) and 0.92 when computed on participant-mean item profiles. The mean inter-item correlation across ratings was 0.40 (range, 0.06–0.80), consistent with a coherent and non-redundant item set. The mean 12-item scale score was 4.43 (SD, 0.45; median, 4.50; range, 2.83–5.00), indicating generally favorable ratings with limited use of the lower response range. Among the nine participants who rated two or more conversations, the ICC(1) was 0.45, with a within-participant standard deviation of 0.33 across chats, suggesting that scores reflected substantial variation both between raters and between conversations rated by the same person, and that the instrument registered chat-to-chat differences in quality rather than fixed rater preferences. When the harm item was included, alpha for the full 13-item set was 0.87 (95% CI, 0.81–0.92).

### Item-level performance

Item means, standard deviations, corrected item–total correlations, and alpha-if-item-deleted for the 12-item scale, computed across all 46 ratings, are shown in Table 3. Corrected item–total correlations ranged from 0.42 (empathy; language and reading level) to 0.72 (completeness), all exceeding the conventional 0.30 threshold for item retention. No item’s deletion would meaningfully increase alpha (maximum alpha-if-deleted, 0.88), supporting retention of all 12 items.

**Table 3.** Item Statistics for the 12-Item Patient-AI Rating Quality Scale.

| **Item** | **Mean** | **SD** | **Corrected item–**<br>**total r** | **Alpha if**<br>**deleted** |
| --- | --- | --- | --- | --- |
| 1. Understandable | 4.52 | 0.51 | 0.53 | 0.88 |
| 2. Useful | 4.52 | 0.66 | 0.64 | 0.87 |
| 3. Language and reading level | 4.57 | 0.62 | 0.42 | 0.88 |
| 4. Respectful | 4.80 | 0.40 | 0.46 | 0.88 |
| 5. Empathic | 4.24 | 0.71 | 0.42 | 0.88 |
| 6. Complete | 4.24 | 0.71 | 0.72 | 0.86 |
| 7. Accurate | 4.43 | 0.66 | 0.61 | 0.87 |
| 9. Personalized | 4.17 | 1.08 | 0.66 | 0.87 |
| 10. Focused on issues that matter most | 4.33 | 0.60 | 0.68 | 0.87 |
| 11. Helped understand my health | 4.39 | 0.68 | 0.68 | 0.87 |
| 12. Prepared for future care | 4.52 | 0.62 | 0.62 | 0.87 |
| 13. Informed to make future decisions | 4.39 | 0.65 | 0.65 | 0.87 |

Abbreviation: PAIR, Patient-AI Rating. Statistics are based on 46 ratings from 14 participants. Item 8 (harm) is reported separately. Item numbering follows the instrument.

### Inter-item covariance structure

The full inter-item covariance matrix is shown in Table 4 (mean inter-item covariance, 0.18). Items 9–13 — personalization, focus on patient priorities, understanding health, preparation for health encounters, and informed decision support — covaried strongly with one another, suggesting a cluster reflecting the value of the AI communication for patient engagement practices. Completeness (item 6) and accuracy (item 7) showed substantial covariance both with each other and with the items 9–13 cluster. Finally, the personalization item exhibited the largest variance of any item (1.17, vs 0.16–0.50 for all others).

**Table 4.** Inter-item Covariance Matrix for the Patient-AI Rating Quality Scale.

| Item | 1 | 2 | 3 | 4 | 5 | 6 | 7 | 9 | 10 | 11 | 12 | 13 |
| --- | --- | --- | --- | --- | --- | --- | --- | --- | --- | --- | --- | --- |
| 1. Understandable | 0.26 | 0.17 | 0.21 | 0.10 | 0.12 | 0.12 | 0.08 | 0.20 | 0.12 | 0.10 | 0.08 | 0.06 |
| 2. Useful | 0.17 | 0.43 | 0.16 | 0.10 | 0.16 | 0.23 | 0.21 | 0.26 | 0.20 | 0.17 | 0.21 | 0.19 |
| 3. Language/level | 0.21 | 0.16 | 0.38 | 0.11 | 0.08 | 0.11 | 0.06 | 0.19 | 0.10 | 0.15 | 0.08 | 0.06 |
| 4. Respectful | 0.10 | 0.10 | 0.11 | 0.16 | 0.05 | 0.09 | 0.06 | 0.12 | 0.06 | 0.10 | 0.08 | 0.06 |
| 5. Empathic | 0.12 | 0.16 | 0.08 | 0.05 | 0.50 | 0.25 | 0.14 | 0.38 | 0.10 | 0.10 | 0.03 | 0.08 |
| 6. Complete | 0.12 | 0.23 | 0.11 | 0.09 | 0.25 | 0.50 | 0.27 | 0.49 | 0.23 | 0.24 | 0.18 | 0.24 |
| 7. Accurate | 0.08 | 0.21 | 0.06 | 0.06 | 0.14 | 0.27 | 0.43 | 0.37 | 0.17 | 0.18 | 0.21 | 0.20 |
| 9. Personalized | 0.20 | 0.26 | 0.19 | 0.12 | 0.38 | 0.49 | 0.37 | 1.17 | 0.34 | 0.35 | 0.26 | 0.31 |
| 10. Focused on issues that matter most | 0.12 | 0.20 | 0.10 | 0.06 | 0.10 | 0.23 | 0.17 | 0.34 | 0.36 | 0.27 | 0.18 | 0.22 |
| 11. Understand health | 0.10 | 0.17 | 0.15 | 0.10 | 0.10 | 0.24 | 0.18 | 0.35 | 0.27 | 0.47 | 0.28 | 0.31 |
| 12. Prepared for future care | 0.08 | 0.21 | 0.08 | 0.08 | 0.03 | 0.18 | 0.21 | 0.26 | 0.18 | 0.28 | 0.39 | 0.32 |
| 13. Informed to make future health decisions | 0.06 | 0.19 | 0.06 | 0.06 | 0.08 | 0.24 | 0.20 | 0.31 | 0.22 | 0.31 | 0.32 | 0.42 |

Abbreviation: PAIR, Patient-AI Rating. Based on 46 ratings from 14 participants. Item 8 (harm) is excluded from the quality scale.

### Harm item

Consistent with its design as a safety screen, our harm item had a pronounced floor effect: harm was reported as “Not at all” on 44 of 46 ratings (96%), “A little” on one, and “Mostly” on one. At the participant level, 2 of 14 participants reported any degree of harm on at least one rated conversation; the single “Mostly” rating came from a participant who rated eight conversations and reported no harm on the other seven, consistent with a harm experience specific to one conversation rather than a general negative rater disposition. The item’s corrected correlation with the 12-item quality total was 0.09, and its inclusion did not improve scale reliability, supporting its treatment as a construct distinct from patient-reported communication quality. The restricted variance and low item–total correlation are expected properties of a sentinel item in a low-risk sample and do not indicate item failure.

## Discussion

In this initial patient-partnered development of a tool to rate the quality of patient-AI health conversations, the 12-item PAIR quality scale demonstrated good internal consistency, with a Cronbach’s alpha of 0.88 that remained robust when accounting for clustering of the data (0.85 when restricted to one rating per participant; 0.92 on participant-mean profiles). All corrected item–total correlations exceeded conventional retention thresholds, meaning that no item’s removal would meaningfully improve reliability, and the mean inter-item correlation of 0.40 indicated a coherent item set without redundancy. These preliminary findings support the internal consistency of the PAIR instrument as well as feasibility of the rating process: patient advocates were able to retrieve their own prior AI conversations and rate them independently through an anonymous survey, at a median of two conversations per participant. Because the protocol invited participants to rate multiple conversations, this study also provides an early look at whether a patient-reported measure registers differences between AI conversations rather than differences between raters alone. Less than half of the variance in scale scores was attributable to stable between-rater differences (ICC[1], 0.45), with a within-participant standard deviation of 0.33 across chats, meaning that the same participants scored their different conversations differently. An instrument intended to evaluate patient-facing AI information quality must vary with communication quality and not just by rater. These data offer preliminary evidence of that sensitivity. However, this clustering analysis rests on nine participants who rated more than one conversation and the estimate requires confirmation in a larger sample.

Perceived harm was very low in our sample. As it was conceived as a sentinel safety indicator rather than a quality dimension (accuracy/error served this role), it showed the expected floor in this low-risk sample — no harm on 96% of ratings — a near-zero correlation with the quality scale, and no contribution to scale reliability, confirming that harm and patient-AI conversation quality are distinct constructs in these data.

Patient-centered quality assessment of AI conversations through the PAIR tool, as well as through future patient-derived measures, are complementary to, not in competition with, physician-anchored evaluation. Instruments that quantify accuracy, completeness, and documentation quality from the clinician perspective answer questions that are essential for the deployment of healthcare AI tools, including patient-facing AI. PAIR opens the evaluation space to new constructs — the experience of the patient receiving patient-focused communication. Future evaluations of patient-facing AI systems could incorporate both: physician-anchored tests of accuracy and safety, alongside patient-reported measures of understanding, trust, and usability.^9^

The PAIR instrument may be useful for patient-facing AI tool developers, for clinicians and hospitals deploying patient-facing AI tools, and for patients themselves, as they consider aggregate patient-reported performance of a growing number of health AI tools in the field. Finally, the PAIR instrument may provide a tangible way to include patient perspectives in long-term monitoring of operant AI tools and overall organizational AI governance.^21^

## Limitations

This study has important limitations. It was conducted in a small sample of highly engaged patient advocates who are active AI users, which likely contributed to elevated item means (all ≥ 4.17 on a 5-point scale) and a relatively restricted response range; further validation will be necessary with a broader range of patients and languages. While our measure has face validity due to its creation process, these data do not support claims to criterion or construct validity.

## Conclusions

Patients are already conducting millions of consequential but unmeasured health conversations with AI systems daily. Assessing the patient experience of these conversations is an essential part of meaningful measurement currently missing from the field. The PAIR instrument is a preliminary, patient-partnered first step toward filling this gap.

## Supporting information

Supplementary STROBE Checklist

## Data Availability

The de-identified data supporting the findings of this study are available from the corresponding author upon request, with no restrictions on data sharing.

## Article Information

### Reporting guideline

This study is reported in accordance with the STROBE statement for cross-sectional studies; the completed checklist is provided as a supplementary file.

### Data Access and Responsibility

Adam Rodman had full access to all the data in the study and takes responsibility for the integrity of the data and the accuracy of the data analysis.

### AI Assistance

The investigators selected the statistical analyses. Claude Fable 5 (Anthropic) was used to assist in developing the analysis code, which was executed in Python. GPT-6 Astra (OpenAI) was used to assist with manuscript preparation, including editing, formatting, and preparation of the instrument figure and reporting checklist. Human investigators reviewed the AI-assisted code and manuscript output.

Authors take responsibility for the integrity of the data, the accuracy of the data analysis, and the final manuscript content.

### Author Contributions

Conceptualization: S.K.B., A.R., E.L. Methodology: A.R., S.K.B., E.L., E.J.T. Instrument development and patient partnership: E.L., S.K.B. Investigation and data collection: E.L., Y.B., S.K.B. Formal analysis: A.R., Y.B. Writing — original draft: A.R. Writing — review and editing: all authors. T.S., F.C.B., M.N., C.W provided feedback on the manuscript. Supervision: S.K.B., A.R. Funding acquisition: S.K.B., A.R. All authors approved the final manuscript and agreed to its posting as a preprint and submission for journal publication.

