## Supplementary STROBE Checklist for "Development and Preliminary Evaluation of the Patient–Artificial Intelligence Rating Instrument"

### *Cross-sectional study*

### Title, abstract, introduction, and methods

| Item | Recommendation | Manuscript location / reporting |
|---|---|---|
| 1(a) | Indicate the study's design with a commonly used term in the title or the abstract. | p. 2, Abstract: Design identifies a cross-sectional survey. Also stated on pp. 1 and 4. |
| 1(b) | Provide in the abstract an informative and balanced summary of what was done and what was found. | pp. 2–3, Abstract. |
| 2 | Explain the scientific background and rationale for the investigation being reported. | pp. 5–6, Introduction. |
| 3 | State specific objectives, including any prespecified hypotheses. | p. 2, Abstract: Objective; p. 6, final Introduction paragraph. Instrument development and preliminary evaluation are the stated objectives. |
| 4 | Present key elements of study design early in the paper. | p. 2, Abstract: Design; pp. 6–9, Methods: Instrument development, Instrument, and Participants and setting. |
| 5 | Describe the setting, locations, and relevant dates, including periods of recruitment, exposure, follow-up, and data collection. | pp. 8–9, Participants and setting: Insight Panel and patient peers; anonymous online survey; ratings collected June 5–14, 2026. Recruitment dates are not separately specified. |
| 6(a) | Give the eligibility criteria, and the sources and methods of selection of participants. | pp. 8–9, Participants and setting: adults aged ≥18 years with prior LLM use for personal health queries; email recruitment through the Insight Panel and peer contacts. |
| 7 | Clearly define all outcomes, exposures, predictors, potential confounders, and effect modifiers. Give diagnostic criteria, if applicable. | p. 8, Instrument: 12 quality items plus separately scored perceived-harm item; pp. 9–10, Statistical analysis; pp. 20–21, Tables 1–2. Exposure is participants' prior AI conversations (p. 2). No confounder-adjusted or effect-modification analyses. |
| 8* | For each variable of interest, give sources of data and details of methods of assessment (measurement). Describe comparability of assessment methods if there is more than one group. | pp. 8–10, Instrument, Participants and setting, and Statistical analysis; p. 24, Figure 1. Self-administered survey, 5-point response scale, one response per conversation, identifiers linking repeat ratings. Table 1 footnote defines portal use (p. 20). |

### Methods: bias, sample size, and analysis

| Item | Recommendation | Manuscript location / reporting |
|---|---|---|
| 9 | Describe any efforts to address potential sources of bias. | pp. 7–8, cognitive interviews to refine item interpretation; p. 9, anonymous survey; pp. 9–10, analyses addressing repeated ratings; p. 14, selection of engaged AI users and elevated ratings acknowledged. |
| 10 | Explain how the study size was arrived at. | p. 9, Statistical analysis: no formal sample-size calculation; all eligible panel members invited; size determined by panel size and response during the collection window. |
| 11 | Explain how quantitative variables were handled in the analyses. If applicable, describe which groupings were chosen and why. | pp. 8–10, Instrument and Statistical analysis: 1–5 item responses, 12-item quality scale, harm summarized separately, first-rating and participant-mean analyses. Table 1 (p. 20) identifies demographic categories and digital-literacy grouping. |
| 12(a) | Describe all statistical methods, including those used to control for confounding. | pp. 9–10, Statistical analysis: Cronbach α and Feldt confidence intervals; clustering sensitivity analyses; random-effects ANOVA and ICC(1); item statistics, covariance/correlation matrices, and descriptive harm distributions. No confounder-adjusted analyses. |
| 12(b) | Describe any methods used to examine subgroups and interactions. | Not applicable: no subgroup comparisons or interaction tests are reported. Repeat-rater sensitivity analyses are described under item 12(e). |
| 12(c) | Explain how missing data were addressed. | p. 10, Statistical analysis and Sample: complete PAIR ratings only; demographics and AI-use data use available responses; no imputation. Four incomplete survey submissions excluded. Table 1 footnote (p. 20) gives demographic and AI-use denominators. |
| 12(d) | If applicable, describe analytical methods taking account of sampling strategy. | pp. 9–10, Statistical analysis: repeated ratings addressed by first-rating and participant-mean sensitivity analyses and variance partitioning. Convenience sampling is described on pp. 2 and 8–9; no probability-sampling weights are reported. |
| 12(e) | Describe any sensitivity analyses. | p. 10, Statistical analysis: Cronbach α recalculated using each participant's first chronological rating and participant-mean item profiles. Results on p. 11. |

### Results

| Item | Recommendation | Manuscript location / reporting |
|---|---|---|
| 13(a)* | Report numbers of individuals at each stage of study, eg numbers potentially eligible, examined for eligibility, confirmed eligible, included in the study, completing follow-up, and analysed. | p. 10, Sample: 14 participants contributed 46 complete ratings; 4 additional incomplete survey submissions were excluded. Five participants rated one conversation and nine rated multiple conversations. |
| 13(b)* | Give reasons for non-participation at each stage. | p. 10, Sample: incompleteness is the stated reason for excluding 4 survey submissions. |
| 13(c)* | Consider use of a flow diagram. | No flow diagram is provided. |
| 14(a)* | Give characteristics of study participants (eg demographic, clinical, social) and information on exposures and potential confounders. | p. 20, Table 1: participant characteristics; p. 21, Table 2, and p. 11, Use types: conversation purposes and information entered into AI tools. |
| 14(b)* | Indicate number of participants with missing data for each variable of interest. | p. 10, Sample: demographics available for 13 of 14 participants; all 46 analyzed ratings complete on all 13 items. |
| 15* | Report numbers of outcome events or summary measures. | pp. 11–12, Internal consistency, Item-level performance, Inter-item covariance structure, and Harm item; pp. 22–23, Tables 3–4. |
| 16(a) | Give unadjusted estimates and, if applicable, confounder-adjusted estimates and their precision (eg, 95% confidence interval). Make clear which confounders were adjusted for and why they were included. | p. 11: α estimates and 95% CIs for all ratings, first ratings, and the 13-item set; participant-mean α and ICC(1) are point estimates without CIs. Item estimates in Table 3 (p. 22). |
| 16(b) | Report category boundaries when continuous variables were categorized. | p. 20, Table 1: education categories and digital-literacy grouping (agree or strongly agree) are identified. Age is summarized continuously as median and IQR |
| 16(c) | If relevant, consider translating estimates of relative risk into absolute risk for a meaningful time period. | Not applicable: no relative-risk estimates are reported. |
| 17 | Report other analyses done, eg analyses of subgroups and interactions, and sensitivity analyses. | p. 11: first-rating and participant-mean α, repeat-rater variance partitioning, and α including harm; p. 12: participant-level harm distribution and covariance structure; Table 4 (p. 23). |

### Discussion and funding

| Item | Recommendation | Manuscript location / reporting |
| --- | --- | --- |
| 18 | Summarise key results with reference to study objectives. | pp. 12–13, opening Discussion paragraphs. |
| 19 | Discuss limitations of the study, taking into account sources of potential bias or imprecision. Discuss both direction and magnitude of any potential bias. | p. 13: clustering estimate based on only nine repeat raters needs confirmation. p. 14, Limitations: small, highly engaged sample; likely elevated item means and restricted range; no criterion/construct-validity claims. Bias magnitude is not formally quantified. |
| 20 | Give a cautious overall interpretation of results considering objectives, limitations, multiplicity of analyses, results from similar studies, and other relevant evidence. | pp. 12–14, Discussion, Limitations, and Conclusions: preliminary internal consistency and conversation-level variation; complementary role alongside clinician evaluation; broader validation needed. |
| 21 | Discuss the generalisability (external validity) of the study results. | p. 14, Limitations: engaged AI-using patient advocates, restricted response range, and need for broader patient and language representation. |
| 22 | Give the source of funding and the role of the funders for the present study and, if applicable, for the original study on which the present article is based. | pp. 14–15, Article Information: Funding/Support and Role of the Funder/Sponsor. NLM/NIH award R01LM014689; no funder role in study conduct, reporting, or submission decision. |

* Where applicable, report exposed and unexposed groups separately. No such comparison is reported in this study. Cohort- and case-control-only checklist items are not applicable to this cross-sectional design.

STROBE: Strengthening the Reporting of Observational Studies in Epidemiology. Checklist recommendations: www.strobe-statement.org.
